# Telemedicine and Patient Satisfaction in Saudi Arabia

**DOI:** 10.1101/2021.06.22.21259347

**Authors:** Amjad Alfaleh, Abdullah Alkattan, Mohammed Salah, Mona Almutairi, Khlood Sagor, Alaa Alageel, Khaled Alabdulkareem

**Affiliations:** General Directorate of Medical Consultations, Ministry of Health, Riyadh, Saudi Arabia; General Directorate of Primary Health Centers, Ministry of Health, Riyadh, Saudi Arabia; Assisstant Deputy Minister for Primary Healthcare, Ministry of Health, Riyadh, Saudi Arabia

**Keywords:** Telemedicine, Healthcare system, Saudi Arabia, Ministry of health

## Abstract

**Objectives:** Patients’ satisfaction with the healthcare system is a good indicator for measuring the quality of health services. This study aims to determine patients’ satisfaction with different types of telemedicine services (937 Call Center and Sehha Application) provided by the Ministry of Health in Saudi Arabia.

**Methods:** A cross-section study was conducted to evaluate consumers’ satisfaction toward telemedicine services in Saudi Arabia. A systematic random sampling method was used to collect consumers from each of the two telemedicine services including 937 medical call center and the Sehha application.

**Results:** Two hundred and forty-nine (249) randomly chosen consumers of 937 medical call center and the Sehha application have been answered the predesigned questionnaire about satisfaction towards different items of the two medical services. Among 249 consumers of telemedicine services, 83.14% of them were satisfied in general with medical services compared to 8.03% of consumers who were not satisfied. The satisfaction percentages toward physicians’ recommendations, communication skills, listening skills, and waiting time were 77.29%, 83.53%, 85.14%, and 67.87% respectively.

**Conclusion:** Telemedicine applications are commonly used nowadays in most developed countries and some developing countries in order to maximize the delivery of healthcare to patients with different medical conditions. The overall satisfaction rates toward different telemedicine services in Saudi Arabia were high, and there was no significant difference in concern to the satisfaction rates between 937 medical call center and Sehha application. In general, consumers of telemedicine services were satisfied, and most of them considered advising other people to use them.

## Introduction

Healthcare organizations in every country provided health services that include diagnosis of medical conditions, surgical procedures, dispensing medications, and follow-up of patients with chronic diseases.^[1]^ These services are intended to meet people’s needs, ensure good quality of life and decrease incidences of disease.^[2]^ For a healthcare system needed to provide high-quality health care to a specific population; it is important to avail high quality human and material resources, ensure full access to the services including elderly and people living in rural areas, and ensure patients’ compliance and satisfaction.^[3]^ Patients’ satisfaction toward the healthcare system is a good indicator for measuring the quality of health services.^[4]^ The level of satisfaction reflects on issues related to medical practice and can be obtained through patient surveys.^[5]^

Telemedicine applications are commonly used nowadays in most of the developed countries and some developing countries in order to maximize the delivery of healthcare to patients with different medical conditions.^[6]^ These applications, implemented with the best human and material resources, are designed to measure the vital signs, take symptoms and patient history, make possible diagnosis and treatment.^[7]^

In the United States of America (USA), telemedicine applications are commonly used and more than 90% of patients believe that telehealth has many advantages.^[8]^ Previous studies revealed that telemedicine interventions reduce patients’ overload in hospitals, improve clinical outcomes and increase patients’ satisfaction.^[9]^ However, the greatest drawback of telemedicine in the USA from patients’ perspective are the costs and availability.^[10]^ One of the most effective telemedicine services used in the USA is the poison Control Call Center (PCC) which provides toxicological consultations for both healthcare professionals and non-professionals people.^[11]^ PCC recruits well-trained pharmacists and nurses to give precise and instant telephone-based toxicological consultations.^[12]^

A previous study was done in Egypt about patient satisfaction with medical services in primary healthcare centers (PHCs). The study revealed that the level of patient satisfaction positively correlated with the quality of the service provided. The level of patient satisfaction was low in PHCs that provided poor medical services while it was high in those with good medical services.^[13]^ About two-third of studies done related to patient satisfaction have some limitations because of the lack of comparability and investigated only a few dimensions of satisfaction of medical services. Such limitations may lead to misleading results and could be the reason for the high percentage of patient satisfaction obtained in some trials, especially those studies related to telemedicine.^[14]^

The Ministry of Health (MOH) in the Kingdom of Saudi Arabia (KSA) introduced telemedicine services in 1433 H. These services are provided through 937 Medical Call Center, Seha Mobile Application. These services are aiming to provide access to patients unable to visit primary health care clinics due to living far away from cities and help to reduce costs consumed without any significant needs.

This study aims to determine patients’ satisfaction with different types of telemedicine services (937 Call Center and Sehha Application) provided by the Ministry of Health in Saudi Arabia.

## Subject and Methods

### Study design and patients’ recruitment

A cross-section study was conducted during the period from May to October 2020 to evaluate consumers’ satisfaction toward telemedicine services in Saudi Arabia. A systematic random sampling method with a sample interval of 1:13 was used to collect consumers from each of the two telemedicine services including 937 medical call center and Sehha application to evaluate and compare consumers’ satisfaction along with the two services. The calculated sample size was 245 consumers according to the equation n= z2*(pq)/d2*design effect, where p is estimated at 91% (based on Saudi MOH references found at https://www.moh.gov.sa), z=2.576 equivalent to confidence level of 99%, d= 5% and design effect of (1). Two hundred forty-nine (249) consumers agreed to participate in the survey. A predesigned questionnaire was collected from the consumers through telephone and short message service (SMS). Experts reviewed the questionnaire for validity and reliability of all items.^[15]^ The questionnaire included closed questions regarding socio-demographic characteristics, type of services, and data regarding satisfaction towards different items of the services. The open-ended question was used to determine consumers’ suggestions for improvement of the services.

Inclusion criteria included any consumer aged 18 years and above who accept to participate in the study. Exclusion criteria included consumers with a non-medical chief complaint and those with any health problem that limits their participation in the study.

### Statistical Analysis

Data were analyzed using Microsoft Excel, 2013 (Microsoft Corp., Redmond, Washington USA). Chi-squared test and student’s t-test were used to compare between the two services regarding satisfaction and socio-demographic variables. The level of significance was considered at a p-value <0.05.

### Ethical consideration

Informed consent was taken from the participants after an explanation about the study. The information taken from the consumers is kept confidential and not used for other purposes than the study. Those who refused to participate in the study were excluded. The study proposal was reviewed and approved by an ethics review committee (the Central Institutional review board committee) in the Saudi Ministry of Health. The approval letter for this study was given in June 2020 with the central IRB log number: 20-91M.

## Results

Two hundred and forty-nine (249) randomly chosen consumers of 937 medical call center and the Sehha application have been answered the predesigned questionnaire about satisfaction towards different items of the two medical services. The mean age of the total 249 consumers included in this study was 34.9 ± 9.89 years, and 50.2% of them were male. The majority of the consumers were Saudi citizens (93.59%), and approximately 45% of them were living in the Riyadh region (see figure.1). Detailed consumers’ baseline characteristics are shown in table.1. Among 249 consumers of telemedicine services in Saudi Arabia, 83.14% of them were satisfied in general with medical services provided by the ministry of health compared to 8.03% of the consumers who were not satisfied (p-value < 0.001). The satisfaction percentages toward physicians’ recommendations, communication skills, listening skills, and waiting time were 77.29%, 83.53%, 85.14%, and 67.87% respectively (see figure.2).

**Figure.1.**
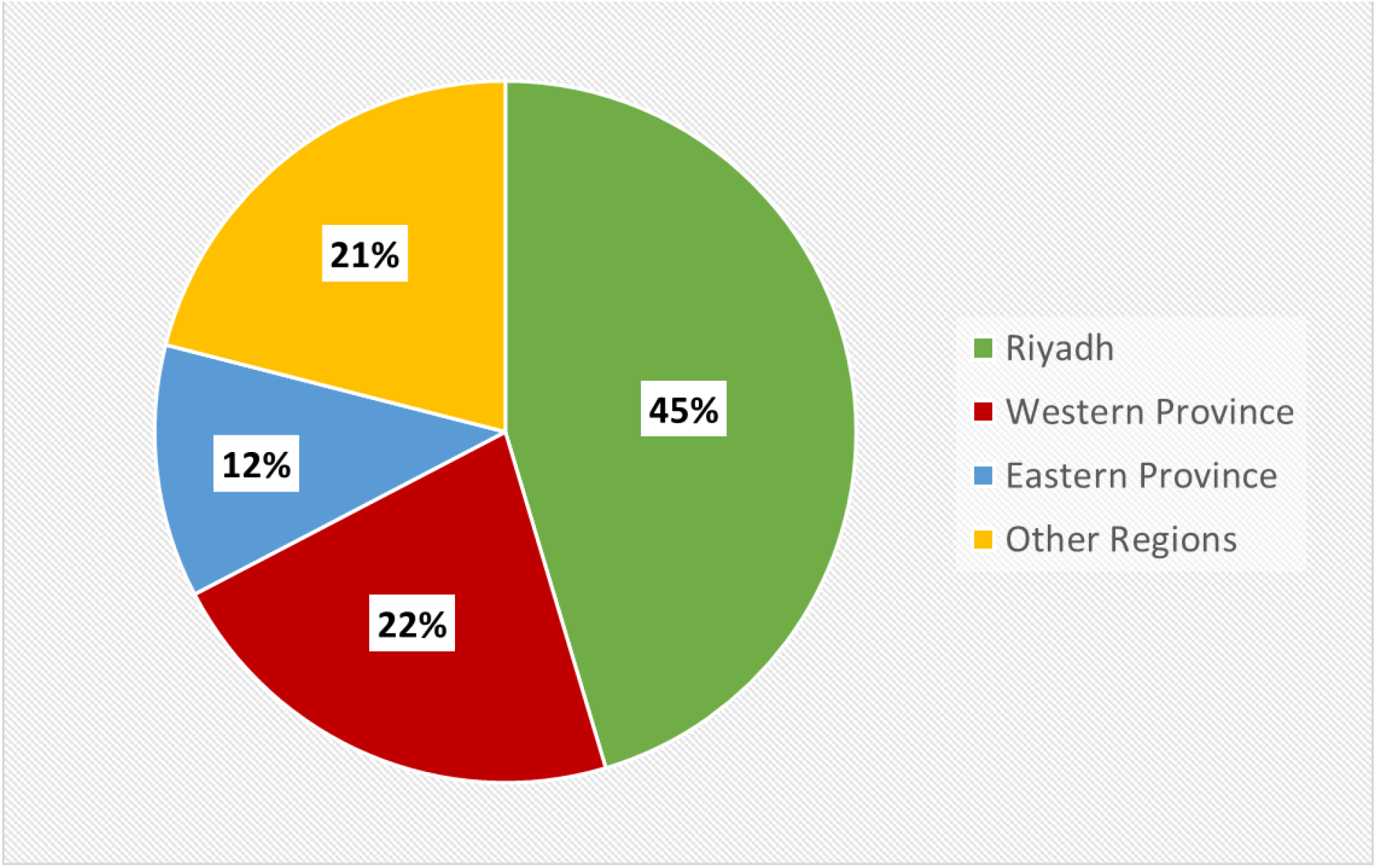
Distribution of Telemedicine’s Consumers in Saudi Arabia.

**Table.1.** Consumers’ Baseline Characteristics.

| Variables | All telemedicine consumers (N=249) | 937 medical call center's consumers (N=203) | Sehha application's consumers (N=46) |
| --- | --- | --- | --- |
| Mean age (in years) | 34.90 | 34.90 | 34.96 |
| Ages younger than 40 years (%) | 72.23 | 71.81 | 73.91 |
| Ages between 40-60 years (%) | 25.21 | 25.00 | 26.09 |
| 60 years or older (%) | 2.56 | 3.19 | 0 |
| Saudi nationality (%) | 93.59 | 94.15 | 91.3 |
| Living Region |  |  |  |
| Riyadh (%) | 45.38 | 45.30 | 45.65 |
| Western province (%) | 21.96 | 22.65 | 26.09 |
| Eastern province (%) | 11.65 | 10.35 | 17.40 |
| Other Regions (%) | 21.01 | 21.70 | 10.86 |

**Figure.2.**
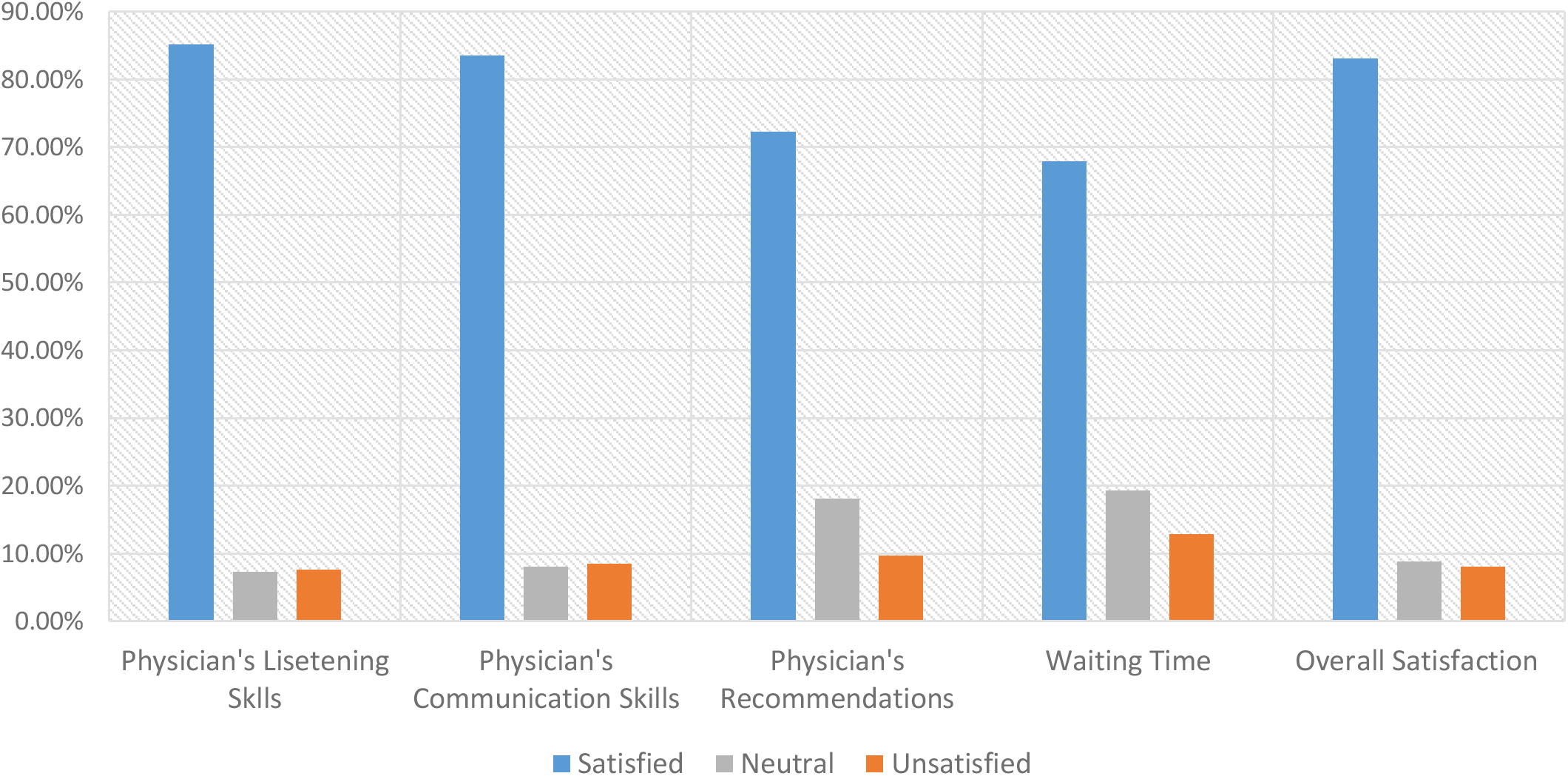
Consumers’ Satisfaction Rates Toward Telemedicine Services.

Regarding 937 medical call center’ consumers, the mean age was 34.9 ± 10 years, and 53.2% of them were male. The general Satisfaction percentage concerning 937 medical call center’ service among consumers was 83.74% compared to 8.86% were unsatisfied (p-value < 0.001). The satisfaction percentages concerning the medical call center physicians’ recommendations, communication skills, listening skills, and waiting time were 73.89%, 83.25%, 82.75%, and 73.89% respectively (see figure.3). On the other hand, the Sehha application’s 46 consumers included in this study mean age was 34.96 ± 9.5 years, and 36.96% of them were male. The general satisfaction percentage with respect to Sehha application’s medical services among its consumers was 80.44% compared to 4.35% who were not satisfied (p-value < 0.001). Sehha application’s satisfaction percentages about physicians’ recommendations, communication skills, listening skills, and waiting time were 65.21%, 84.78%, 95.65%, and 56.51% respectively (see figure.4 and table.2). Neutral responses from consumers regarding general medical services of 937 medical call center and Sehha application were 7.39% and 15.22% respectively.

**Figure.3.**
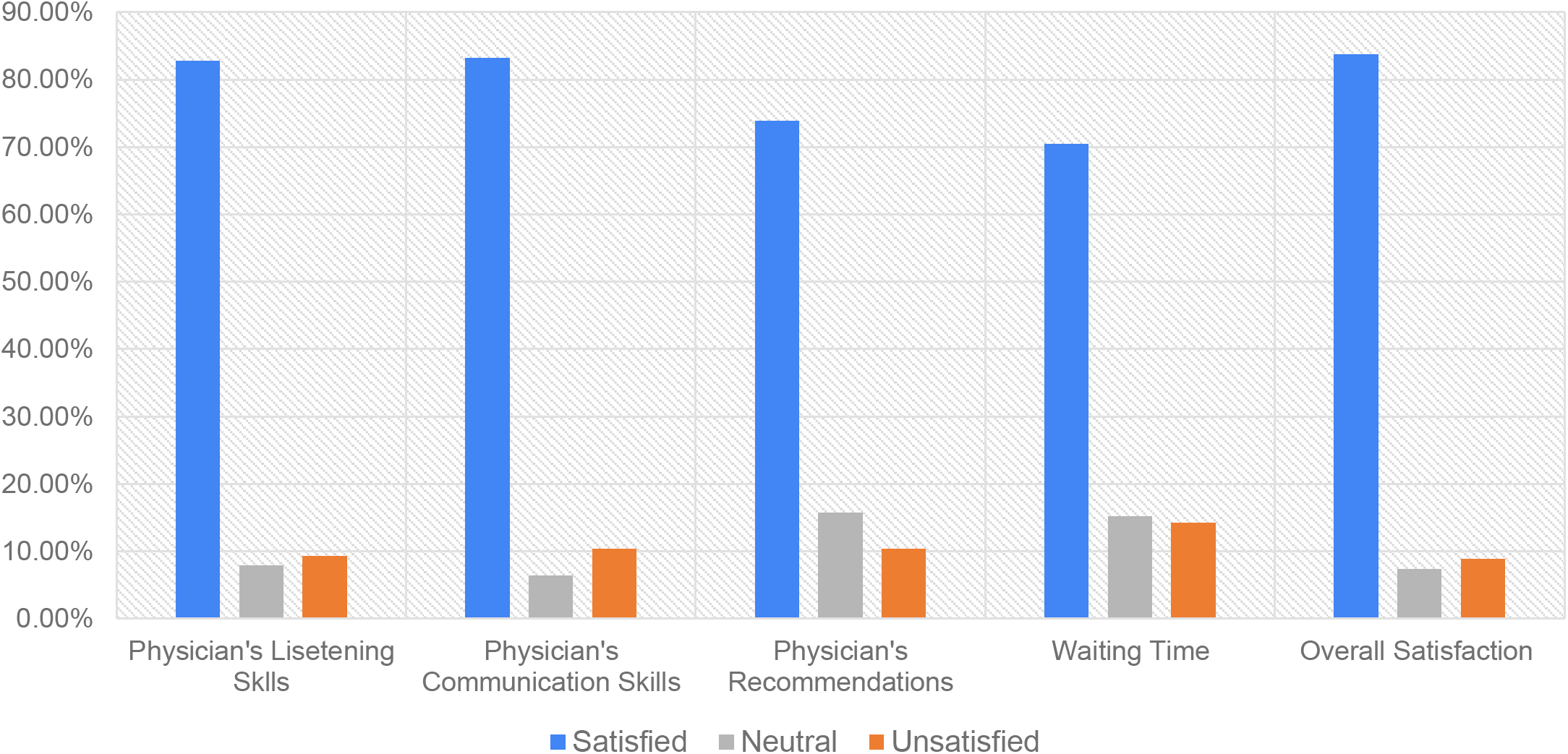
Consumers’ Satisfaction Rates Toward 937 Medical Call Center.

**Figure.4.**
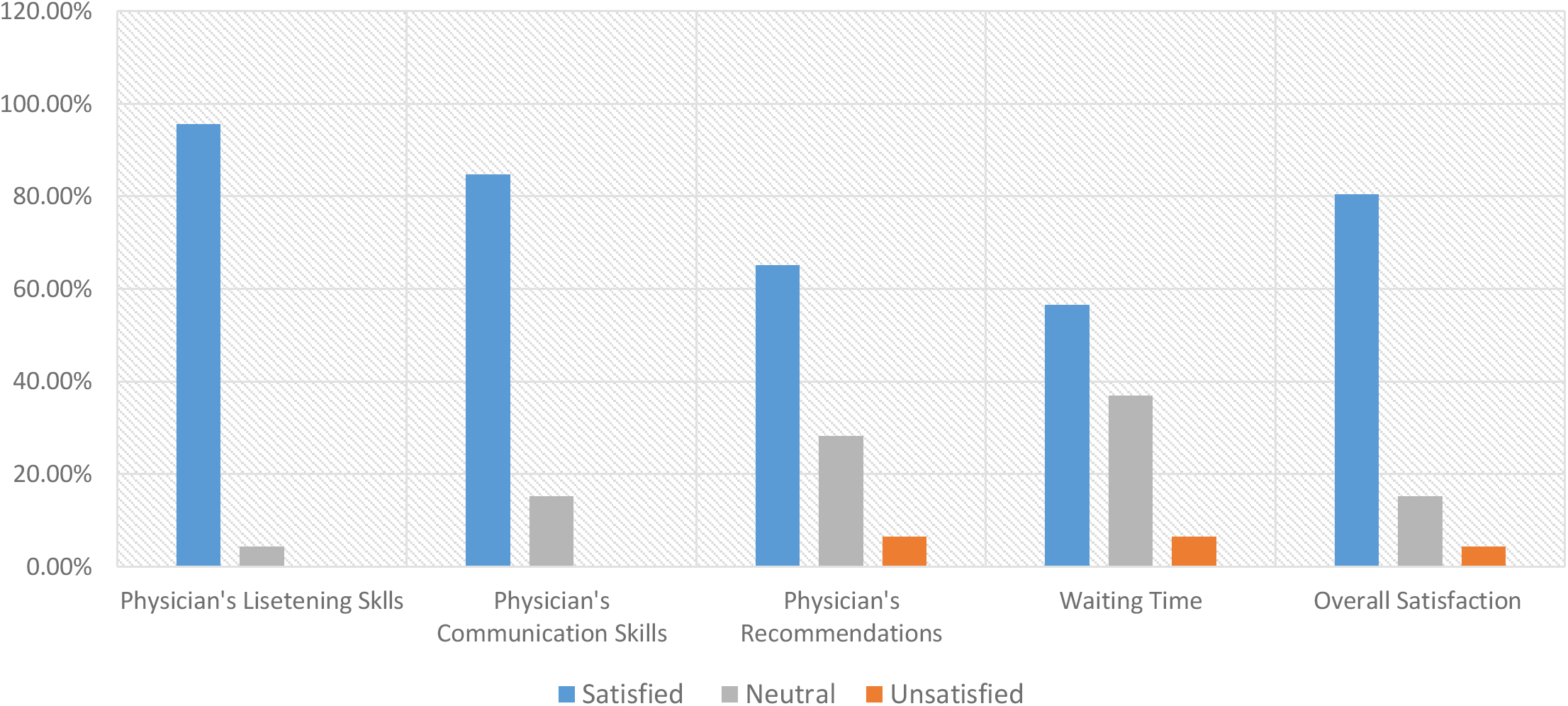
Consumers’ Satisfaction Rates Toward Sehha Application.

More than 93% of 249 participants in this study considered advising others to use the Saudi MOH telemedicine services, and more than 74% of the participants had used these telemedicine services more than once (see figure.5 and figure.6). Concerning on each telemedicine service, 75.5% of 937 medical call center’ consumers considered advising others to take medical consultations and recommendations from this service, and 77.37% of them were calling more than once. However, 100% of Sehha application’s consumers take into consideration to advise others to use it, and 84.78% of them used Sehha application more than once (see table.2).

**Figure.5.**
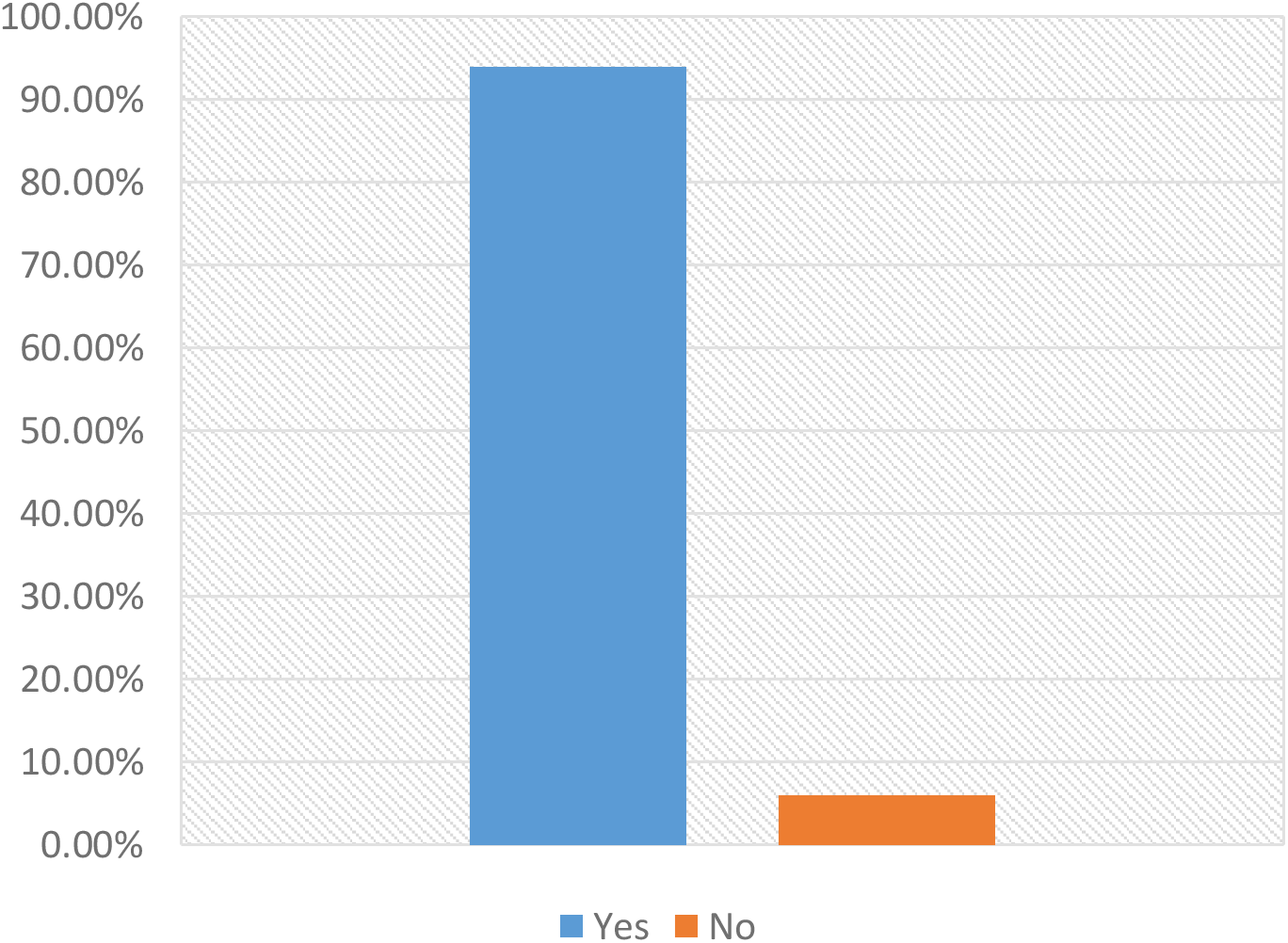
Consider to Advice Others to Use Telemedicine.

**Figure.6.**
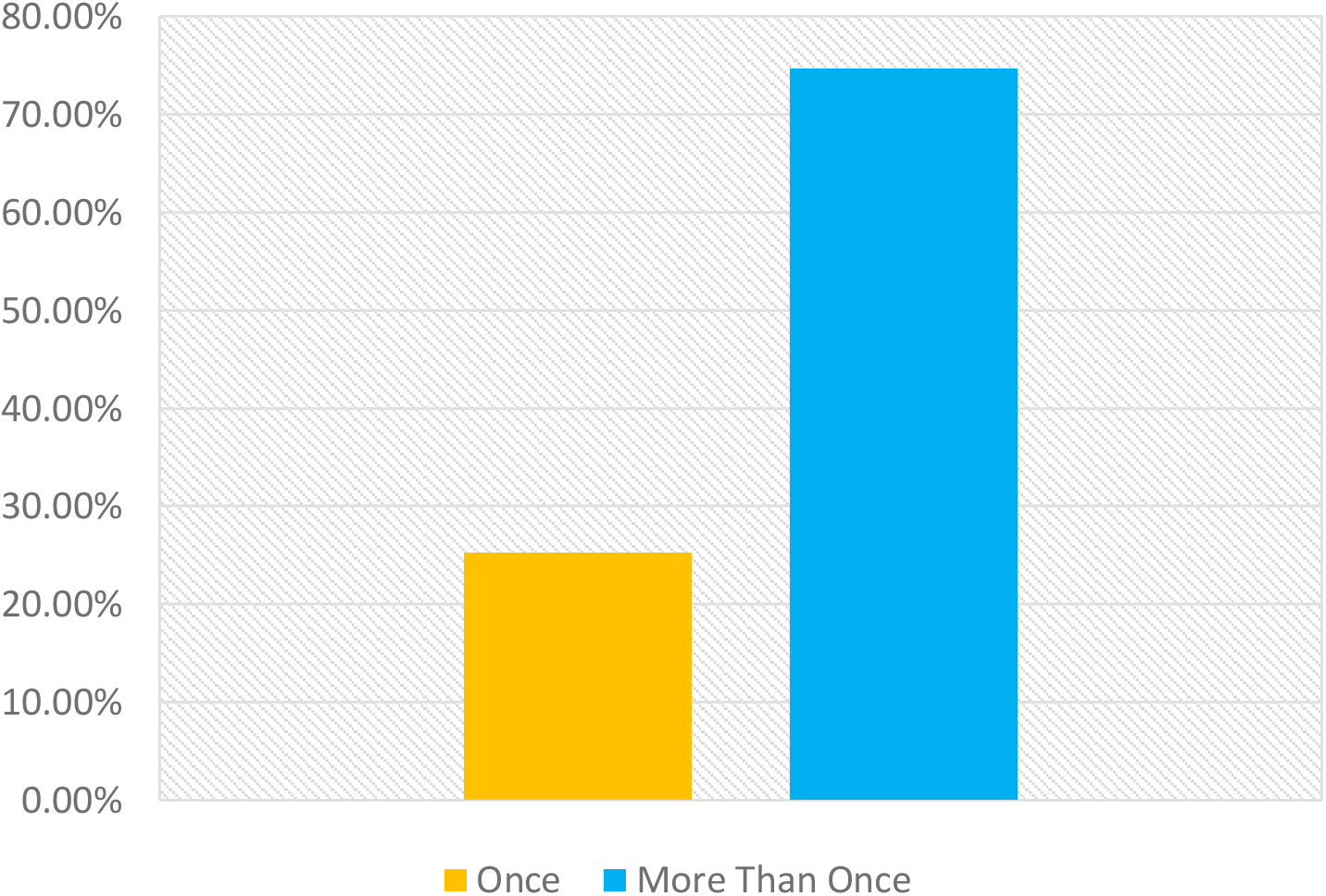
Frequency of Using Telemedicine.

**Table.2.** Percentages of Consumers’ Satisfaction toward Telemedicine Services and Frequency of Use.

| Variables | All telemedicine consumers (N=249) | 937 medical call center's consumers (N=203) | Sehha application's consumers (N=46) | p-value |
| --- | --- | --- | --- | --- |
| Physicians' recommendations (%) | 72.29 | 73.89 | 65.21 | 0.49 |
| Physicians' communication skills (%) | 83.53 | 83.25 | 84.78 | 0.93 |
| Physicians' listening skills (%) | 85.14 | 82.75 | 95.65 | 0.53 |
| Waiting time (%) | 67.87 | 70.44 | 56.51 | 0.41 |
| Overall medical service (%) | 83.14 | 83.74 | 80.44 | 0.86 |
| Used telemedicine more than once (%) | 74.7 | 77.37 | 84.78 | 0.51 |
| Advising others to use telemedicine (%) | 93.98 | 75.5 | 100 | 0.74 |

## Discussion

The concept of telemedicine first appeared in the Lancet journal in 1879 after a few years of inventing the telephone.^[16]^ This concept about telemedicine was figured in the cover of an American magazine in 1925, as this figure showed a physician at his clinic diagnosing a patient lying at home.^[17]^ After years, and until today, telephone, radio, and video communications between healthcare providers and patients are still observed.^[18]^ The consumers’ satisfaction toward telemedicine services was evaluated in different published studies in the 21^st^ century. These previous studies found that patients were highly satisfied after being discussed about possible diagnosis and treatment options through video or telephone contact.^[19,20,21,22]^

In this study, the consumers’ satisfaction toward telemedicine services provided by the Saudi MOH was estimated. The results showed high satisfaction rates regarding physicians’ recommendations, communication skills, listening skills, waiting time, and overall medical services. Besides, there were no significant differences in the satisfaction rates regarding various concerns between 937 medical call center and the Sehha application (p-values > 0.05). Regarding the telemedicine consumers’ ages, the results revealed that 72.23% of the consumers were younger than 40 years old compared to 25.21% of them were middle-aged (p-value < 0.001). The satisfaction percentages towards waiting time and physicians’ recommendations of all telemedicine services were not high enough (not exceeds 74%), however, there were no significant differences when compared to other satisfactions rates related to physicians’ communication skills, listening skills, and overall medical services (p-value > 0.05). The lower satisfaction rates for waiting time could be due to the overload of consultations related COVID-19 pandemic.

The most common comments from the consumers were complaining about the waiting time to talk with a physician and also complaining about not having a direct extension to talk with a female physician. The last complaint frequently noted in Saudi Arabia, since some Saudi women refused to talk or be medically examined concerning obstetrics and gynecological consultations by male physicians.^[23]^

Several previous studies were evaluating the satisfaction of patients toward telemedicine services provided by various medical institutions globally. Five trials revealed that patients were highly satisfied after applying telemedical consultations for them. Moreover, many of the patients included in some of these studies were preferring the use of telemedicine over conventional visits.^[24,25,26,27,28]^ These findings were almost similar to this current study in Saudi Arabia and showing more advantages of using telemedicine not only for seeking patients’ satisfaction but also for reducing hospital readmission and emergency overload.

### Limitations and bias

Limitations of the study are related to the cross-section design including recall bias and a self-reported questionnaire. Also, the sample may not cover the entire spectrum of the general population, however, the researchers have done efforts to avoid selection bias, where only a few of the non-respondents reflected their opinion. The vast majority of them did not respond as they did not have time to participate in the survey, and few of them were not satisfied with the service.

### Conclusion

The overall satisfaction rates toward different telemedicine services in Saudi Arabia were high despite not enough satisfaction rates observed regarding waiting time because of consultations’ overload during the COVID-19 pandemic. There was no significant difference in concern to the satisfaction rates between 937 medical call center and Sehha application. In general, consumers of telemedicine services provided by the Saudi MOH were satisfied, and most of them think that these services deserved to be used again and considered to advise other people to use them.

## Data Availability

N/A

## Study’s Questionnaire

Study's Questionnaire
| Question | Answer |
| --- | --- |
| -age |  |
| -Gender | -male - female |
| -Nationality | -Saudi - non-Saudi |
| Region | --Riyadh - Eastern region - Ahsaa - norther boarders - Quryiat - Jouf - Tabuk - Hail - Qaseem - Madina - Mekka - Jeddah - Qunfotha -Taif - Aseer - Bisha - Najran - Jezan - Baha |
| Is this the first to use the service? | - Yes – No |
| Are you satisfied with time of waiting until physician answer? | Strongly satisfied – Satisfied – okay – not satisfied – strongly not satisfied |
| Are you satisfied with how physician deals with you? | Strongly satisfied – Satisfied – okay – not satisfied – strongly not satisfied |
| Did the physician listen to your complain and give you enough time to explain it? | Strongly satisfied – Satisfied – okay – not satisfied – strongly not satisfied |
| Are you satisfied with physician's advices and recommendations? | Strongly satisfied – Satisfied – okay – not satisfied – strongly not satisfied |
| Are you Satisfied in general about this telemedicine service? | Strongly satisfied – Satisfied – okay – not satisfied – strongly not satisfied |
| Will you advice other people to call this service? | Yes - No |
| Do you have any recommendations to develop this service> | open question |

